# Quantitative fibrosis features from digitized H&E-stained liver biopsies reveal granular insights into fibrosis and disease progression in metabolic dysfunction-associated steatohepatitis: a retrospective analysis of STELLAR-3 and STELLAR-4

**DOI:** 10.1101/2025.06.12.25328580

**Authors:** Ylaine Gerardin, Adam Stanford-Moore, Neel Patel, Yibo Zhang, Jun Zhang, Pratik Mistry, Deeksha Kartik, Yi Liu, Nicholas Indorf, Darren Fahy, Geetika Singh, Jonathan Glickman, Murray Resnick, Lily Windholz, Naim Alkhouri, Andrew Billin, Tim Watkins, Jacqueline Brosnan-Cashman, Christina Jayson, Justin Lee, Ben Glass, Andrew H. Beck, Janani Iyer, Michael G. Drage, Lara Murray, Robert Egger

## Abstract

**Background:** Histologic staging of metabolic dysfunction-associated steatohepatitis (MASH) requires semiquantitative assessment of hepatocellular ballooning, steatosis, lobular inflammation, and fibrosis and is fraught with pathologist variability. We hypothesize that quantitative histologic analysis will better reflect the continuous distribution of histologic features, and thus the disease biology.

**Methods:** Using LiverExplore, a suite of machine learning models that detect and classify liver tissue regions, lobular zones, cell types, and fibrosis subtypes from hematoxylin and eosin-stained whole slide images, we characterized the MASH disease microenvironment in participants of the STELLAR-3 and STELLAR-4 trials (NCT04052516). The correlation of model-derived human interpretable features (HIFs) with pathologist-provided MASH CRN grades and fibrosis stages, AI-generated continuous CRN grades and stages, non-invasive biomarkers, transcriptomics, and patient outcomes was assessed.

**Results:** Model-quantified steatosis, lobular inflammation, hepatocellular ballooning, and fibrosis features were significantly correlated with pathologist CRN grades/stages, while model-derived tissue and cell features revealed quantitative changes in the disease microenvironment as MASH progressed. Pathological and advanced fibrosis HIFs were correlated with non-invasive metrics of fibrosis and a gene signature associated with hepatic stellate cells. HIFs associated with nodular or advanced fibrosis and inflammation were associated with an increased risk of liver-related events in patients from STELLAR-3 and STELLAR-4.

**Conclusions:** AI-powered quantitative characterization of the liver microenvironment delivers context relevant to MASH progression beyond the resolution afforded by categorical CRN scoring, highlighting the promise of this approach for broad applications in drug development.

## Background

Liver fibrosis is a wound healing response to chronic injury and inflammation, manifesting as the excess deposition of collagen and other extracellular matrix components. The development of fibrosis accompanies chronic liver diseases, broadly (1), with specific fibrosis patterns developing in each disorder. The presence of advanced fibrosis (stages F3-F4) has been identified as the most important prognostic indicator of outcomes in longitudinal studies of patients with metabolic dysfunction-associated steatohepatitis (MASH) (2), and the risk of both liver-specific and all-cause mortality increases with fibrosis stage (3). Indeed, patients progressing to cirrhosis have an elevated risk of hepatic decompensation, hepatocellular carcinoma, and cardiovascular events (4,5).

While advanced fibrosis was traditionally thought to be irreversible, recent evidence has shown the ability of fibrosis to regress, even in the setting of cirrhotic MASH (6). While resmetirom and semaglutide have recently been approved by the United States Food and Drug Administration (FDA) for patients with MASH with stage F2-3 fibrosis, there remains no approved therapeutic for patients with compensated cirrhosis (F4 disease). Indeed, despite promising early-phase studies, Phase 3 trials of selonsertib in patients with F3 (STELLAR-3) and F4 MASH (STELLAR-4) failed to meet their primary endpoint (7). Recent results from the Phase 2b SYMMETRY trial have demonstrated significant fibrosis improvement in patients with F4 MASH treated with efruxifermin, a FGF21 analog, for 96 weeks (8). Still, the lack of an approved treatment option for patients with F4 MASH based on a successful phase 3 trial in this population leaves an urgent unmet clinical need.

Manual pathologist assessment of liver histology, particularly fibrosis, is not without limitations. To evaluate fibrosis, pathologists have traditionally relied on special staining [e.g., Masson trichrome (MT) or picrosirius Red (9)] or specialized imaging of adjacent unstained sections (10), rather than using tissue routinely stained with hematoxylin and eosin (H&E), making it difficult to assess fibrosis alongside other histologic features relevant to disease assessment. The gold standard for fibrosis assessment in MASH remains the histologic staging of MT-stained liver biopsies using the MASH clinical research network (CRN) scoring scale, increasing in severity through stages F3 (bridging fibrosis) and F4 (cirrhosis) (11). However, the scoring criteria defined by these guidelines and used by pathologists are semi-quantitative in nature. These shortcomings have led to a high degree of variability in how pathologists interpret fibrosis present within liver biopsies. Furthermore, patients with MASH present across a complex and multifaceted spectrum that can be challenging to capture with manual pathologist review, traditionally reliant on semi-quantitative, categorical staging of fibrosis and other disease components (11–13). Thus, opportunities remain for more in-depth understanding of disease-specific liver microarchitecture alterations.

As pathology workflows become increasingly digital in nature, an opportunity exists to leverage artificial intelligence (AI)-powered digital pathology (DP) algorithms to overcome these challenges. These algorithms have the ability to provide unbiased, quantitative, and reproducible evaluation of whole slide images (WSIs) of histology specimens. Notably, algorithms have been developed to measure the MASH clinical research network (CRN) criteria (hepatocellular ballooning, steatosis, lobular inflammation, and fibrosis), including qFIBS (14) and AIM-MASH (15). Both algorithms have been demonstrated to improve inter-pathologist concordance for scoring fibrosis and recapitulate manually-scored fibrosis clinical trial endpoints, while AIM-MASH has been shown to improve pathologist reproducibility and corroborate manual endpoints for the additional CRN criteria, leading to its qualification by the European Medicines Agency and the FDA (15–18). However, both tools necessitate additional slides than H&E-stained biopsies for fibrosis detection, either unstained (qFIBS) (14) or MT-stained (AIM-MASH) (15). The ability to assess and quantify fibrosis on a H&E-stained slide could streamline liver pathology workflows for fibrosis evaluation, and the integration of such an approach with additional measures of the liver microarchitecture has unique potential to yield critical insights about the fibrotic milieu in liver diseases, such as MASH.

In this study, we conducted a retrospective analysis of STELLAR-3 and STELLAR-4 using an AI-powered DP tool [LiverExplore (For Research Use Only. Not for use in diagnostic procedures)] that comprehensively quantifies features of the liver microarchitecture – including fibrosis – from H&E-stained liver biopsies. Our objectives were to 1) characterize changes in the liver microenvironment that occur as advanced fibrosis progresses and 2) identify histologic features associated with liver-related outcome events in patients with F3 and F4 disease.

## Methods

### Ethics

This study was conducted in accordance with the Declaration of Helsinki and Good Clinical Practice guidelines. All patients provided informed consent for future research and tissue histology, and approval by central institutional review boards was granted for each clinical trial (7,18).

### Datasets

Baseline and post-treatment liver biopsies from two completed MASH clinical trials of selonsertib, STELLAR-3 and STELLAR-4 (NCT04052516) (7), were stained with H&E and MT. Slides were imaged using Aperio AT2 (Leica Biosystems, Deer Park, IL, USA) scanning microscopes to generate digital whole slide images (WSIs). Gene expression (19), NIT metrics, and clinical outcomes were also available for all patients in the dataset.

### AI-mediated liver microarchitecture assessment

LiverExplore (PathAI, Boston, MA) was used to predict tissue regions, cell types, lobular zones, and fibrosis subtypes in evaluable regions of H&E-stained liver biopsy WSIs. Model predictions are summarized in Figure 1. Tissue regions predicted by the model include hepatocellular ballooning, lobular inflammation, steatosis, and normal hepatocyte regions. Cell nuclei were exhaustively classified as plasma cells, lymphocytes, neutrophils, eosinophils, macrophages, fibroblasts, steatotic hepatocytes, normal hepatocytes, ballooned hepatocytes or other cells. Portal tracts, central veins, and the three lobular zones were identified. Fibrosis was detected and subtyped into structural collagen, portal collagen, perisinusoidal fibrosis, periportal fibrosis, perivenular fibrosis, incomplete septal fibrosis, complete septal fibrosis, nodular fibrosis.

**Figure 1.**
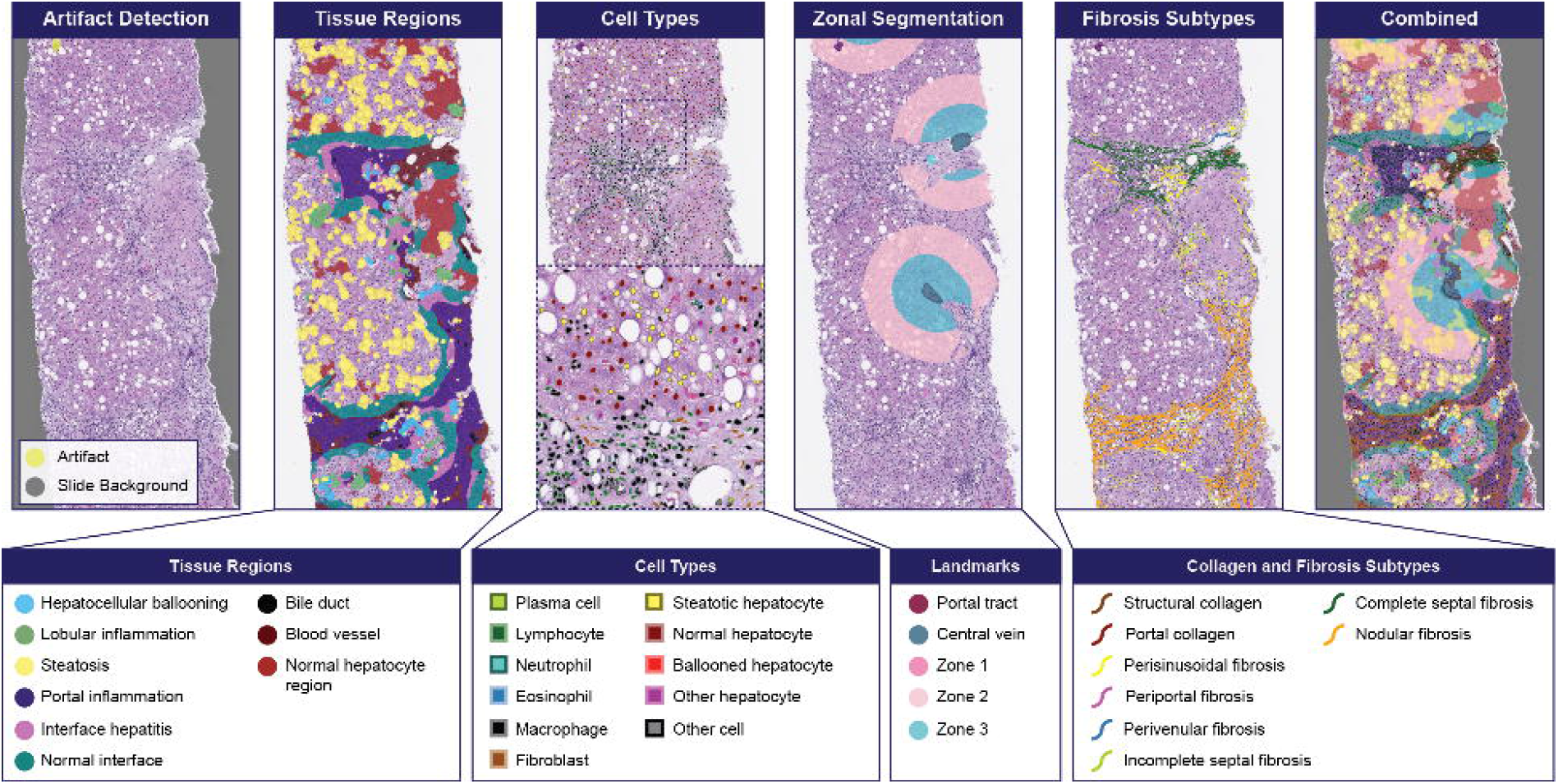
LiverExplore overlays and detected classes.

Accuracy of model predictions was evaluated using nested pairwise analysis (20) to compare outputs with pathologist annotations of cell types (**Supplementary Table S1**), lobular zones (**Supplementary Table S2**), and fibrosis subtypes (**Supplementary Table S3**) in a separate evaluation dataset of samples from a phase 2 clinical trial of icosabutate (NCT04052516) (18).

### Model deployment and feature extraction

WSIs with <4 mm^2^tissue, <1000 total hepatocytes, and <200 hepatocytes per mm^2^tissue were excluded, resulting in N=3728 WSIs across STELLAR-3 and STELLAR-4 (7). All model overlays were used to generate human interpretable features, or HIFs (N>1,000) which describe the liver architecture through the quantification of tissue regions, cell types, zonal regions, and fibrosis subtypes using the process previously described (21). Briefly, features were calculated through the combination of heatmap overlays, yielding relative areas of zonal regions or fibrosis types, relative cell densities, cell density ratios, and substances within a certain distance of others. Continuous CRN fibrosis scores were derived using (7,22) AIM-MASH+ (15) deployed on paired MT slides.

### Expression of transcriptomic modules

Gene expression data for baseline and post-treatment biopsies in the STELLAR-3 and STELLAR-4 datasets were processed into log-normalized counts per million as previously described (19), then used to calculate the hepatic stellate cells-2 (HSC-2) score based on the published list of genes and corresponding weights (23).

### Correlation of AI-predicted histology with outcomes

LiverExplore features in baseline biopsies were independently tested for association with liver-related events (LRE) in STELLAR-3 and in STELLAR-4. Features with >3% missing values across the baseline sample set or relating to artifact or background areas, nuclear morphology features, as well as raw areas and raw counts were excluded. Area proportion and count proportion features were logit-transformed after imputing zeros to half the minimal non-zero value and imputing ones to the average between 1 and the maximal non-one value. All other features were log-transformed after imputing zeros to the half-minimal value. The entire feature set was then normalized using the robust Z-score method.

Cox proportional hazards models for each transformed baseline feature in each data set were fitted to LRE data (N=637 for STELLAR-3; N=643 for STELLAR-4), including progression to cirrhosis for STELLAR-3, with treatment arms as covariates. Of LREs noted in STELLAR-3 (N=94), N=93 (98.9%) were progression to cirrhosis, while N=1 (1.1%) was qualification for liver transplantation (MELD score ≥ 15). The LREs in STELLAR-4 (N=20) were due to ascites (N=9; 45%), hepatic encephalopathy (N=6; 30%), portal hypertension-related upper gastrointestinal bleeding (N=2; 10%), qualification for liver transplantation (MELD score ≥ 15; N=2; 10%), and liver transplantation (N=1; 5%). Samples with missing feature values were excluded. A significance threshold of p=0.05 after false discovery rate correction was used.

## Results

### Association between AI-derived histological features and CRN scores

To understand how AI-predicted histologic features correlate with established MASH histology scoring, the relationship between HIFs and CRN grades/stages was assessed in specimens from STELLAR-3 and STELLAR-4. We identified a HIF quantifying the relevant histology of each CRN component and assessed the distributions of these features across pathologist-assessed grades/stages (**Figure 2**). Features demonstrated strong correlation to manual scores, indicating that this approach accurately captures histologic features relevant for clinical trials.

**Figure 2.**
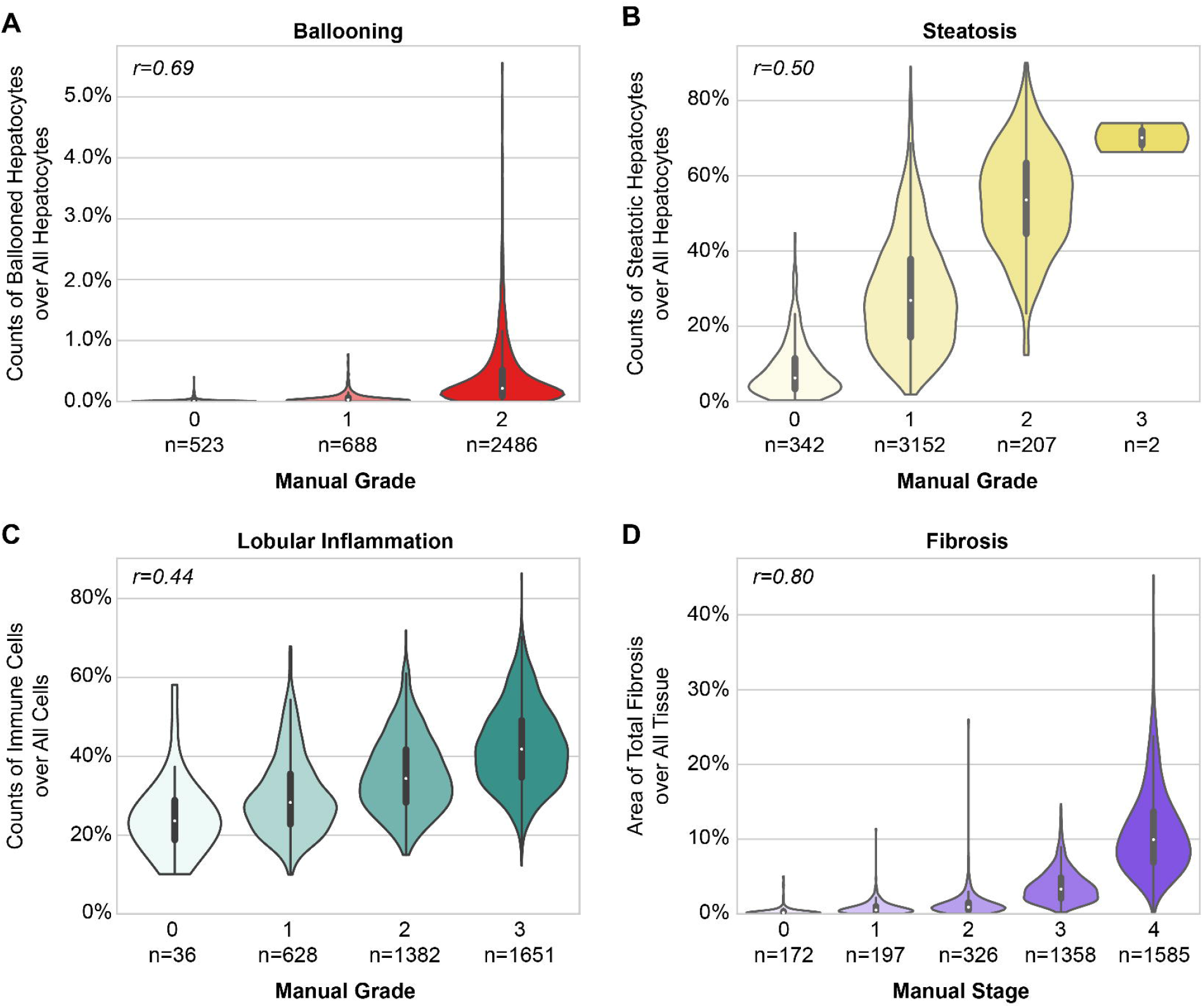
Distribution of LiverExplore-quantified histological substances within pathologist-assessed MASH CRN grades and fibrosis stages. Spearman correlation coefficients are shown.

Given the unique importance of fibrosis for determining MASH severity, we sought to ascertain the changes in liver composition that accompany fibrosis progression. To do so, tissue, cell, and fibrosis HIFs were quantified in WSIs from STELLAR-3 and STELLAR-4 and compared to AI-predicted continuous fibrosis stages (15). The proportionate area of normal hepatocyte regions, representing the amount of healthy tissue, decreased as a function of fibrosis stage (**Figure 3A**), consistent with the notion that healthy tissue decreases as disease progresses. Interestingly, at continuous fibrosis scores above 3.0, large expansions of portal inflammation, blood vessels (**Figure 3A**), nodular fibrosis (**Figure 3B**), immune cell density, and fibroblast density were observed (**Figure 3C**), progressively increasing with fibrosis severity. Intermediate disease stages were characterized by elevated complete septal (bridging) fibrosis, incomplete septal fibrosis, and perisinusoidal fibrosis, which appeared first in early-stage disease with maximum percentages occurring near continuous scores of 4.0. Steatosis tissue and steatotic hepatocyte density both peaked in biopsies with continuous fibrosis scores between 1.0 and 3.0 (**Figure 3A,C**).

**Figure 3.**
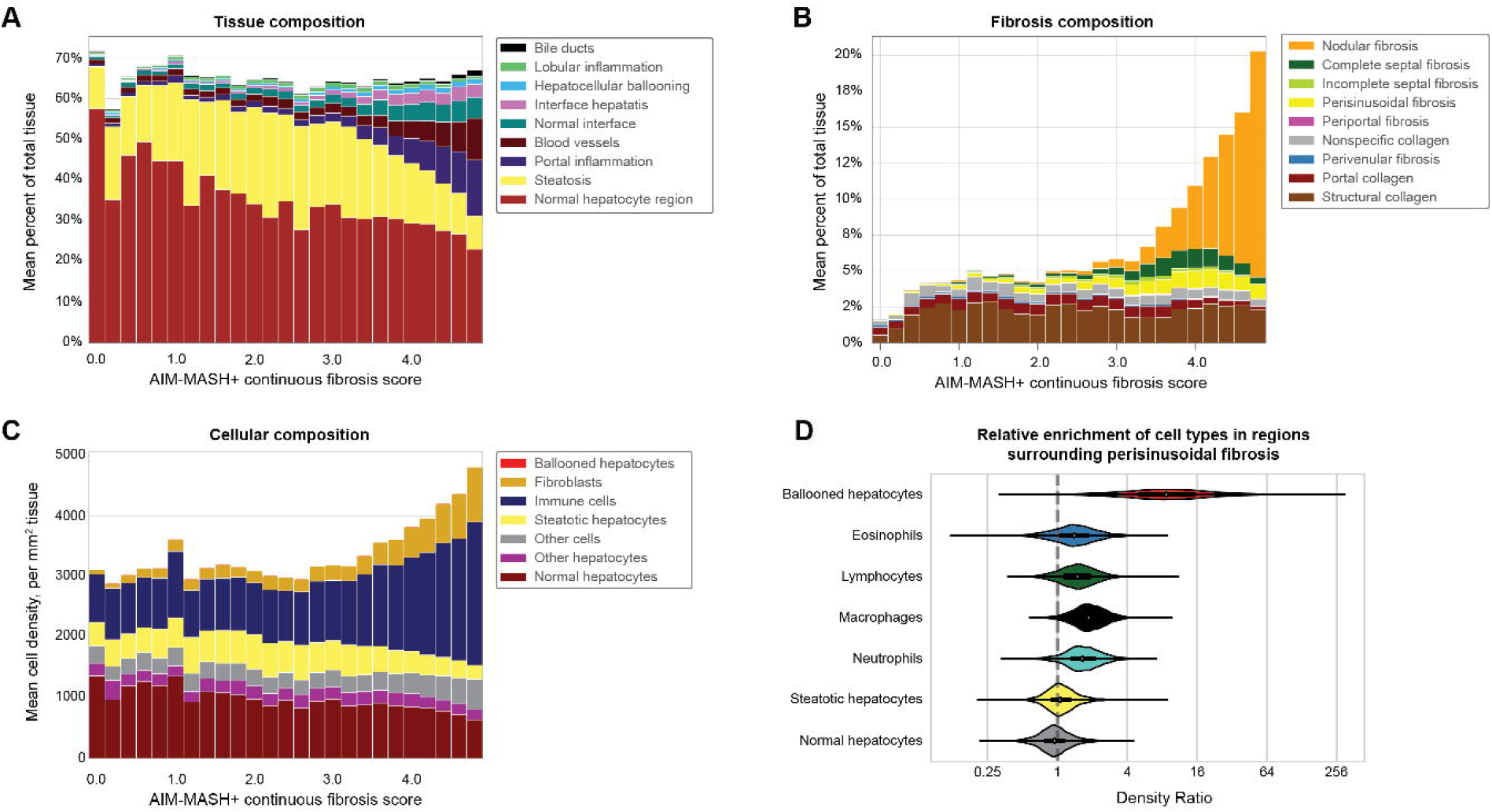
Granular changes in MASH liver tissue composition revealed at sub-ordinal and spatially differentiated resolution with increasing disease severity. Continuous fibrosis scores were binned into 5 equal-sized intervals per integer score, and bars show feature values averaged across all samples falling within each interval for **A)** predicted tissue types, **B)** predicted fibrosis subtypes, and **C)** predicted cell types. **D)** Relative enrichment of each cell type in regions surrounding perisinusoidal fibrosis. Enrichment for each biopsy image is defined by the ratio of cell density within 40 µm of perisinusoidal fibrosis divided by cell density in tissue excluding those proximal regions.

### Evaluation of the cellular composition of specific fibrotic milieu

The identification of discrete fibrosis subtypes in conjunction with other components of the liver parenchyma, such as cell types, may inform a better understanding of the biology associated with each subtype. To address this question, we sought to determine the distribution of cell types in regions predicted to harbor individual fibrosis subtypes. Interestingly, tissue regions surrounding perisinusoidal fibrosis were strongly enriched for ballooned hepatocytes (median of 8.69x density compared to other tissue regions) and moderately enriched for infiltrating immune cells (macrophages=1.87x, neutrophils=1.64x, lymphocytes=1.49x and eosinophils=1.39x density ratio; **Figure 3D**). However, steatotic hepatocytes and normal hepatocytes showed little to no spatial bias towards fibrosis regions. These results suggest that the enrichment of ballooned hepatocytes and certain inflammatory cells near perisinusoidal fibrosis is due to a cell-type specific mechanism.

### Association of HIFs with non-invasive MASH biomarkers

NITs and blood-based biomarkers are widely used for diagnosis (24–26) and assessment of MASH severity (27), as they assess fibrosis (FIB-4, ELF, VCTE, NFS, APRI), inflammation (C-reactive protein [CRP]), cell death (CK-18-M30, and -M65), liver enzyme levels (aspartate transaminase [AST] and alanine transaminase [ALT]), blood triglyceride, and cholesterol levels. To better understand how these metrics associate with liver histology, we performed correlation analysis of HIFs with available NITs from patients enrolled in STELLAR-3 and STELLAR-4 as additional validation (**Figure 4A**).

**Figure 4.**
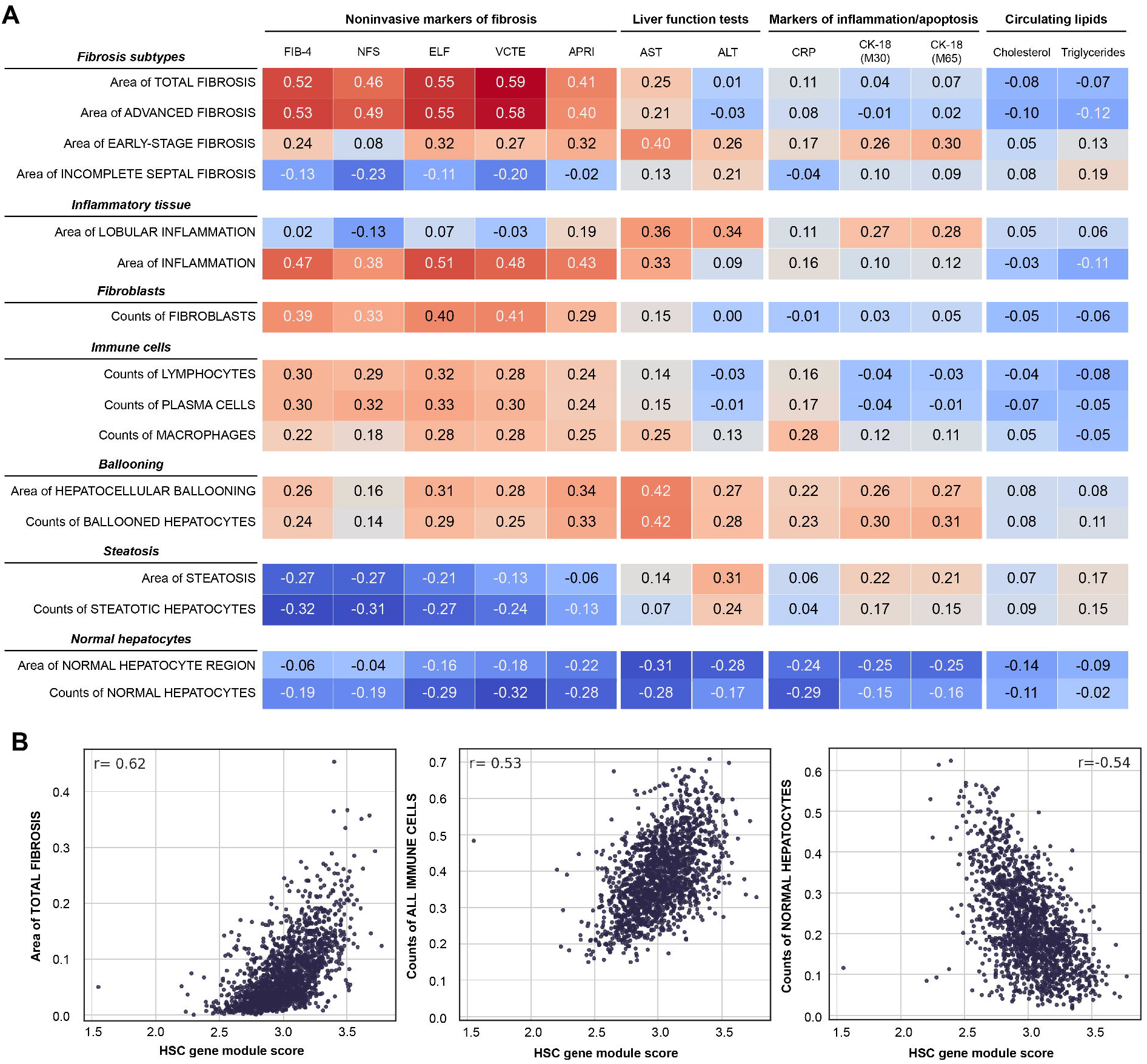
Correlation of LiverExplore features with orthogonal data modalities. **A)** LiverExplore features representing MASH-relevant disease features correlated against blood-based biomarkers of liver function and metabolism (Spearman r). ‘Area’ features are proportionate areas out of all evaluable tissue; ‘count’ features are cell proportions out of all cells. Data includes baseline and post-treatment time points from both studies. **B)** LiverExplore features correlated with gene expression signature of hepatic stellate cells (Spearman r).

Comparisons of fibrosis NITs with fibrosis HIFs revealed strong positive associations between all fibrosis NITs and model-derived total fraction of pathological fibrosis area. Additionally, these NITs were more strongly correlated with the fraction of advanced fibrosis area than with early-stage fibrosis area, potentially indicating a higher sensitivity of fibrosis NITs for advanced fibrosis and cirrhosis (28). Similarly, Aspartate Transaminase Platelet Ratio Index (APRI), which detects advanced fibrosis and cirrhosis, was more strongly correlated with advanced than early-stage fibrosis. At the cellular level, fibrosis NITs correlated positively with the fraction of fibroblasts and negatively with normal hepatocyte area and counts. The association of LiverExplore fibrosis HIFs with fibrosis NITs provides orthogonal validation of model-predicted features.

Having observed increased portal inflammation and immune cells with increasing fibrosis score (**Figure 3A**), we also sought to confirm this association using NITs. Indeed, the fraction of inflammation area in tissue was strongly correlated with fibrosis NITs, suggesting that inflammation could accompany advanced fibrosis development (29).

Of the liver function tests, AST demonstrated stronger correlations with most HIFs than ALT, including positive correlations with early-stage fibrosis, hepatocellular ballooning area, and ballooned hepatocyte counts. CK-18-M30 and CK-18-M65 also demonstrated strong positive correlation with hepatocellular ballooning features (area and cell count) and negative correlation with normal hepatocyte features (area and cell count).

### Association of HIFs with gene expression signatures

Finally, we compared model-derived HIFs with single cell-based transcriptomic modules related to MASH severity (23) (**Figure 4B**). Interestingly, despite LiverExplore not directly quantifying hepatic stellate cells, pooled LiverExplore HIFs by predicted substance were strongly correlated with the hepatic stellate cells-2 (HSC-2) transcriptomic module, representing hepatic stellate cells and containing genes related to collagen and matrix remodeling (23). To further examine this association, we compared individual HIFs to the HSC-2 module score. HSC-2 was positively correlated with the fractions of total pathological fibrosis area and immune cells and negatively correlated with the fraction of normal hepatocytes.

### LiverExplore HIFs are prognostic of MASH progression

Given the changes in model-predicted features accompanying MASH progression, we hypothesized that these features may be prognostic of outcome. To address this question, we tested individual HIFs in baseline biopsies for association with liver-related events (LREs) in STELLAR-3 (7) and STELLAR-4.

We first assessed the contribution of features quantifying tissue regions, cell types, lobular zones, and fibrosis subtypes to LRE risk in STELLAR-3 and STELLAR-4 (**Figure 5A**). Several classes of features demonstrated statistically significant prognostic capabilities in both STELLAR-3 STELLAR-4 (**Figure 5A**; **Supplementary Table X; Supplementary Table Y**). In both trials, features quantifying inflammation were associated with elevated risk of LRE. In particular, the relative area of inflammation was associated with LRE risk in STELLAR-3 (HR 2.37, 95% CI 1.60-3.52; FDR-corrected p<0.0001; **Figure 5C**) and in STELLAR-4 (HR=4.20, 95% CI 1.91-9.22; FDR-corrected p=0.007; **Figure 5D**).

**Figure 5.**
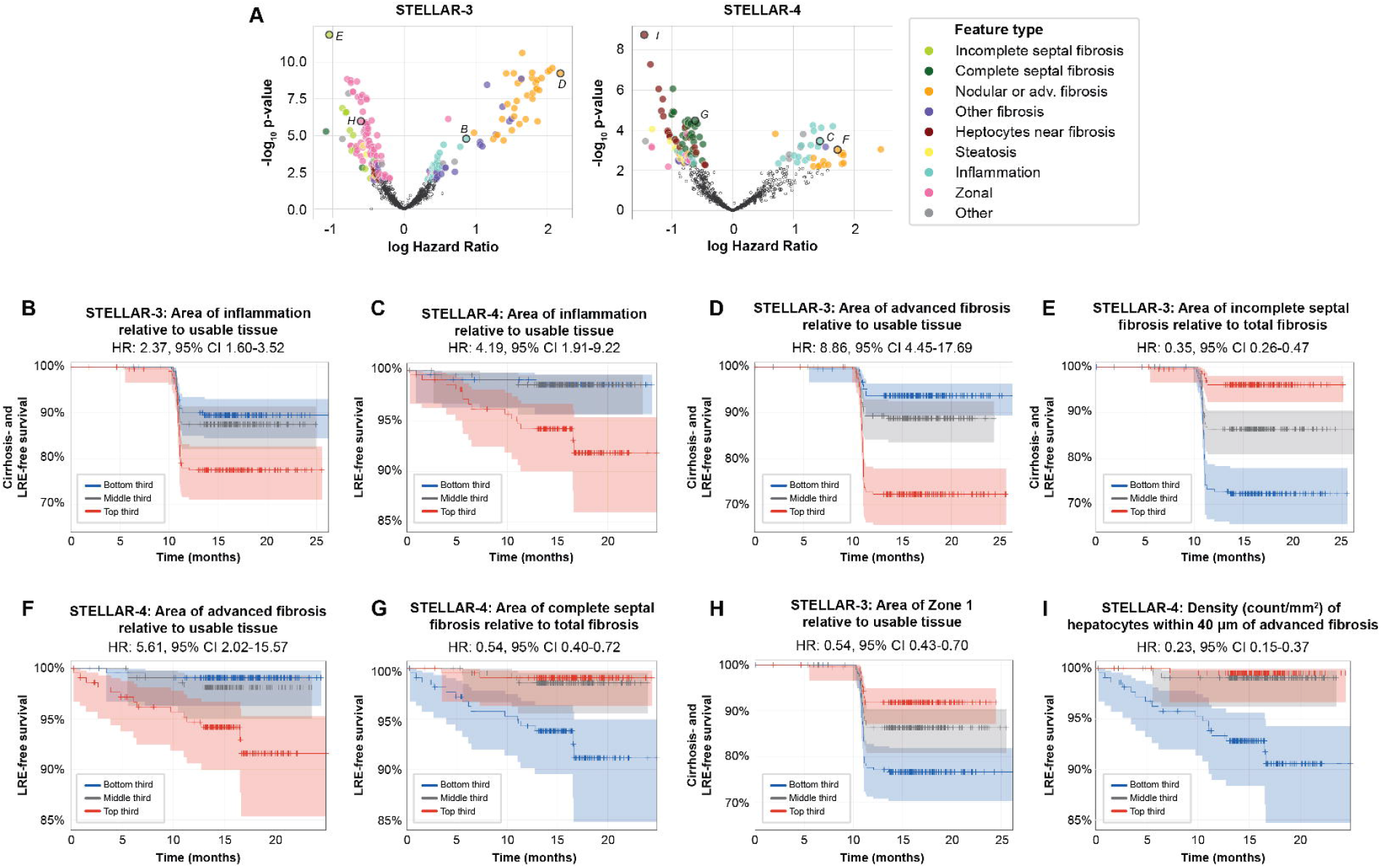
LiverExplore features are prognostic of outcome. **A)** Baseline sample feature value and association with clinical outcomes from STELLAR-3 and STELLAR-4. Each point represents a single feature’s nominal p-value and effect size from Cox regression. Large colored markers indicate features satisfying FDR-adjusted p<0.05, and points outlined in black are highlighted in panels B-I. **B-I**) Cox regression stratifying patients by LiverExplore feature value tertiles to predict cirrhosis- and LRE-free survival in STELLAR-3 (B, D, E, H) and LRE-free survival in STELLAR-4 (C, F, G, I).

Notably, features quantifying and nodular or advanced fibrosis were associated with higher risk of LRE, while features quantifying less severe fibrosis subtypes (incomplete septal in STELLAR-3 and complete septal in STELLAR-4) were associated with lower risk of LRE. In STELLAR-3, elevated areas of advanced fibrosis were associated with worse outcomes (HR=8.86, 95% CI 4.45-17.69; FDR-corrected p<0.0001; **Figure 5D**), while elevated areas of incomplete septal fibrosis were associated with reduced risk of progression (HR=0.35, 95% CI 0.26-0.47; FDR-corrected p<0.0001; **Figure 5E**). Similarly, in STELLAR-4, higher amounts of advanced fibrosis were associated with increased LRE risk (HR=5.61, 95% CI 2.02-15.57; FDR-corrected p=0.01; **Figure 5F**), while higher areas of complete septal fibrosis were associated with reduced LRE risk (HR=0.54, 95% CI 0.40-0.72; FDR-corrected p=0.002; **Figure 5G**). These results suggest that for each study cohort, the dominant fibrosis subtype is directly related to outcomes, with patients with more advanced fibrosis having worse prognosis.

Additional trial-specific features were also identified that were associated with lower LRE risks. In STELLAR-3, features quantifying areas of landmark zones were associated with lower LRE risk (**Figure 5A**), as were features measuring densities of hepatocytes in proximity to fibrosis in STELLAR-4 (**Figure 5A**). Specifically, the relative total area of Zone 1 was associated with reduced LRE risk in STELLAR-3 (HR=0.55, 95% CI 0.43-0.69; FDR-corrected p<0.0001; **Figure 5H**), while the density of total hepatocytes within 40 μm of advanced fibrosis was associated with reduced LRE risk in STELLAR-4 (HR=0.23, 95% CI 0.15-0.37; FDR-corrected p<0.0001; **Figure 5I**). These results are likely driven by the distinct histologies of patients enrolled in each study (e.g., early signs of damage to landmark regions in F3, progression to nodular fibrosis in F4).

## Discussion

The study of advanced fibrosis in MASH has been hindered by variability in pathologist interpretations (due to the need for specialized stains or imaging to score fibrosis), as well as semi-quantitative fibrosis scoring criteria. AI-based DP approaches provide an opportunity to improve our understanding of fibrosis. Here, using a suite of algorithms that provide comprehensive quantification of liver histology (including fibrosis) from an H&E-stained liver biopsy, we characterized a cohort of patients with advanced fibrosis (F3-F4) from the STELLAR-3 and STELLAR-4 trials (7).

The LiverExplore suite of models enables direct quantification of fibrotic composition beyond the categorical CRN stages, providing an unprecedented level of granularity through which fibrosis phenotypes can be studied both individually and in conjunction with other components of the liver microarchitecture. Here, we demonstrated that these quantitative features are aligned with multiple orthogonal metrics of fibrosis severity. The fraction of total fibrosis area within usable tissue was highly correlated with both manual fibrosis stage and non-invasive markers of fibrosis (FIB-4, NFS, ELF, and VCTE). In addition, the assessment of fibrosis subtype areas across AI-derived continuous fibrosis stages revealed that fibrosis subtypes progressively change along a continuum with increasing disease severity, rather than as a stepwise process as reflected in the CRN scoring criteria. Specifically, nodular fibrosis, complete septal fibrosis, and perisinusoidal fibrosis all show distinct patterns of expansion and reduction as fibrosis progresses, thus demonstrating the spectrum of fibrosis present within the categorical fibrosis stages. The quantification of fibrosis progression at the sub-ordinal level underlines the variability present within single fibrosis categories and highlights the value of quantifying the fibrosis continuum in MASH – the potential for a more granular understanding of how fibrosis may progress and, by extension, potentially regress.

Interestingly, while we demonstrated the correlation of model-quantified fibrosis areas with fibrosis NITs, the observed associations were seemingly driven by areas of advanced fibrosis patterns (nodular and complete septal fibrosis), suggesting that fibrosis NITs may lack the sensitivity for detection of early stage fibrosis and incomplete septal fibrosis. As NITs become increasingly used for assessment of fibrosis, further study of the ability of each test to detect individual fibrosis patterns – and specifically regression – is necessary.

Notably, our results demonstrate a clear link between underlying MASH histology and disease outcome. In particular, we observed association between fibrosis subtype features and risk of liver-related events in both STELLAR-3 and STELLAR-4, which enrolled patients with F3 and F4 disease, respectively. If total fibrosis was dominated by advanced subtypes, enrolled patients had a higher risk of LRE in both trials, whereas if less severe fibrosis forms constituted the majority of fibrosis present, patients had lower risk of disease progression. These results are in agreement with previous work demonstrating that FIB-4 can be associated with progression to hepatocellular carcinoma, cirrhosis, and major adverse cardiovascular events (30–32).

Interestingly, features quantifying inflammation were also associated with higher risk of LRE. In this study, we observed several links between inflammation and advanced fibrosis, aligning with previously determined mechanisms of liver fibrogenesis. Hepatic stellate cells (HSCs), when activated by chronic inflammation, generate and deposit excess extracellular matrix components, including collagen, ultimately leading to fibrosis (33). In this study, we observed the fraction of immune cells to expand starting at continuous fibrosis scores of 3.0. Similarly, the model-quantified area of total inflammation showed correlation with fibrosis non-invasive test metrics at levels similar to model-predicted fibrosis area. Lastly, the area of total fibrosis and the fraction of immune cells were both highly correlated with a gene expression signature of HSCs. The association of model-quantified metrics of fibrosis and inflammation with orthogonal metrics of fibrosis and HSC abundance aligns with the biologic link between inflammation, HSC activation, and fibrogenesis.

Taken together, these findings support the notion that the quantitative histologic metrics yielded by LiverExplore can yield granular insights of fibrosis and other features of MASH, with the potential to inform MASH therapeutic development. Information spanning fibrosis subtypes and cell types can be captured from a single H&E-stained image, thereby allowing spatial relationships between these features to be characterized in tandem, an untenable task across separate images (e.g., characterizing cell types and fibrosis from H&E- and MT-stained specimens, respectively), which has the potential to yield novel tissue composition analyses. Furthermore, the observed association between baseline histology and trial outcome supports the concept of histologic biomarkers for precision medicine strategies in MASH. A deeper understanding of the relationship between processes manifesting in distinct histologic events (e.g., inflammation and fibrosis) may enable improved therapeutic targeting strategies, enhance the understanding of the mechanism(s) of action for existing therapeutics, and inform clinical trial design. The ability to more thoroughly detect and quantify histologic changes in fibrosis may lead to a better understanding of the features associated with disease progression and regression and inform future biomarker development for precision medicine or combination therapy approaches in MASH.

A notable strength of this study is the cohort of patients on which the study was performed. The patients were enrolled in the STELLAR-3 and STELLAR-4 clinical trials and had biopsy-confirmed F3 and F4 MASH prior to treatment initiation. As such, the analysis of DP-quantified fibrosis on this patient population is timely and relevant, especially given the lack of approved therapeutics for patients with compensated cirrhosis (F4). As the MASH therapeutic landscape continues to expand, a clear opportunity exists to better understand how the spectrum of fibrosis changes during disease progression in patients with MASH to more effectively treat patients based on their unique fibrotic state. As clinical trials in patients with cirrhosis are increasing in prevalence, the ability to quantify features of fibrosis from a routine H&E-stained biopsy has strong potential to unlock key insights into the liver fibrotic microenvironment in this group of patients. That said, this study is not without limitations. While LiverExplore accurately predicts cell types for which pathologist annotations were collected,, the model cannot identify other cell types, such as hepatic stellate cells, for which it was not possible to collect accurate annotations during development. Other techniques (e.g., RNAseq, spatial transcriptomics) remain necessary for the quantification of these liver cell types, and the integration of these additional modalities has potential to enhance the insights gained from LiverExplore-quantified histology. Furthermore, due to the designs of the STELLAR-3 and STELLAR-4 trials (no placebo arm), we lack the ability to assess the relationship between quantitative histology features and treatment effect. Future studies will address the ability of quantitative histology features to measure and/or predict treatment effect in additional MASH clinical trial cohorts.

In conclusion, we demonstrate the feasibility and utility of quantifying histologic features relating to fibrosis from H&E-stained liver biopsy whole slide images. As the spectrum of therapeutics being investigated in MASH clinical trials expands – including to combination trials and studies of patients with compensated cirrhosis – the approach taken here has the unique potential to dissect therapeutic changes in fibrosis using digitized images of preexisting H&E-stained trial biopsies.

## Data Availability

The histopathology data collected for this study are maintained by PathAI to preserve patient confidentiality and the proprietary image analysis. Any additional information required to reanalyze the data reported in this paper relating directly to the clinical datasets (STELLAR-3, STELLAR-4) will be considered at the discretion of the source institute for the clinical trial in question. Requests will be considered from academic investigators without relevant conflicts of interest for noncommercial use who agree not to distribute the data. Data requests should be sent to Robert Egger.

## Acknowledgements

The authors would like to thank the pathologists who contributed to this study, as well as the software engineering and ML operations teams at PathAI for developing the systems and pipelines used for model development and feature extraction. We are also indebted to the patients enrolled in STELLAR-3 and STELLAR-4 and their families.

## References

1. Parola M, Pinzani M. Liver fibrosis: Pathophysiology, pathogenetic targets and clinical issues. Mol. Aspects Med. 2019;65:37–55.

2. Loomba R, Chalasani N. The hierarchical model of NAFLD: Prognostic significance of histologic features in NASH. Gastroenterology. 2015;149:278–281.

3. Ng CH, Lim WH, Hui Lim GE, Hao Tan DJ, Syn N, Muthiah MD, et al. Mortality outcomes by fibrosis stage in nonalcoholic fatty liver disease: A systematic review and meta-analysis. Clin. Gastroenterol. Hepatol. 2023;21:931–939.e5.

4. Hagström H, Shang Y, Hegmar H, Nasr P. Natural history and progression of metabolic dysfunction-associated steatotic liver disease. Lancet Gastroenterol. Hepatol. 2024;9:944– 956.

5. Sanyal AJ, Van Natta ML, Clark J, Neuschwander-Tetri BA, Diehl A, Dasarathy S, et al. Prospective study of outcomes in adults with nonalcoholic fatty liver disease. N. Engl. J. Med. 2021;385:1559–1569.

6. Sanyal AJ, Anstee QM, Trauner M, Lawitz EJ, Abdelmalek MF, Ding D, et al. Cirrhosis regression is associated with improved clinical outcomes in patients with nonalcoholic steatohepatitis. Hepatology. 2022;75:1235–1246.

7. Harrison SA, Wong VW-S, Okanoue T, Bzowej N, Vuppalanchi R, Younes Z, et al. Selonsertib for patients with bridging fibrosis or compensated cirrhosis due to NASH: Results from randomized phase III STELLAR trials. J. Hepatol. 2020;73:26–39.

8. Noureddin M, Rinella ME, Chalasani N, Neff G, Lucas K, Rodriguez M, et al. GS-012 Efruxifermin improves fibrosis in participants with compensated cirrhosis due to MASH: results of a 96-week, randomized, doubleblind, placebo-controlled, phase 2b trial (SYMMETRY). J. Hepatol. 2025;82:S8–S9.

9. Standish RA, Cholongitas E, Dhillon A, Burroughs AK, Dhillon AP. An appraisal of the histopathological assessment of liver fibrosis. Gut. 2006;55:569–578.

10. Astbury S, Grove JI, Dorward DA, Guha IN, Fallowfield JA, Kendall TJ. Reliable computational quantification of liver fibrosis is compromised by inherent staining variation. J. Pathol. Clin. Res. 2021;7:471–481.

11. Kleiner DE, Brunt EM, Van Natta M, Behling C, Contos MJ, Cummings OW, et al. Design and validation of a histological scoring system for nonalcoholic fatty liver disease. Hepatology. 2005;41:1313–1321.

12. Bedossa P, FLIP Pathology Consortium. Utility and appropriateness of the fatty liver inhibition of progression (FLIP) algorithm and steatosis, activity, and fibrosis (SAF) score in the evaluation of biopsies of nonalcoholic fatty liver disease. Hepatology. 2014;60:565–575.

13. Pai RK, Kleiner DE, Hart J, Adeyi OA, Clouston AD, Behling CA, et al. Standardising the interpretation of liver biopsies in non-alcoholic fatty liver disease clinical trials. Aliment. Pharmacol. Ther. 2019;50:1100–1111.

14. Liu F, Goh GB-B, Tiniakos D, Wee A, Leow W-Q, Zhao J-M, et al. QFIBS: An automated technique for quantitative evaluation of fibrosis, inflammation, ballooning, and steatosis in patients with nonalcoholic steatohepatitis. Hepatology. 2020;71:1953–1966.

15. Iyer JS, Juyal D, Le Q, Shanis Z, Pokkalla H, Pouryahya M, et al. AI-based automation of enrollment criteria and endpoint assessment in clinical trials in liver diseases. Nat. Med. 2024;1–10.

16. Pulaski H, Harrison SA, Mehta SS, Sanyal AJ, Vitali MC, Manigat LC, et al. Clinical validation of an AI-based pathology tool for scoring of metabolic dysfunction-associated steatohepatitis. Nat. Med. 2025;31:315–322.

17. Abdurrachim D, Lek S, Ong CZL, Wong CK, Zhou Y, Wee A, et al. Utility of AI digital pathology as an aid for pathologists scoring fibrosis in MASH. J. Hepatol. 2025;82:898– 908.

18. Harrison SA, Alkhouri N, Ortiz-Lasanta G, Rudraraju M, Tai D, Wack K, et al. A phase IIb randomised-controlled trial of the FFAR1/FFAR4 agonist icosabutate in MASH. J. Hepatol. [Internet]. 2025;Available from: 10.1016/j.jhep.2025.01.032

19. Conway J, Pouryahya M, Gindin Y, Pan DZ, Carrasco-Zevallos OM, Mountain V, et al. Integration of deep learning-based histopathology and transcriptomics reveals key genes associated with fibrogenesis in patients with advanced NASH. Cell Rep Med. 2023;4:101016.

20. Gerardin Y, Shamshoian J, Shen J, Le N, Prezioso J, Abel J, et al. Improved statistical benchmarking of digital pathology models using pairwise frames evaluation [Internet]. arXiv [cs.CV]. 2023;Available from: http://arxiv.org/abs/2306.04709

21. Diao JA, Wang JK, Chui WF, Mountain V, Gullapally SC, Srinivasan R, et al. Human-interpretable image features derived from densely mapped cancer pathology slides predict diverse molecular phenotypes. Nat. Commun. 2021;12:1613.

22. Hirschfield GM, Shiffman ML, Gulamhusein A, Kowdley KV, Vierling JM, Levy C, et al. Seladelpar efficacy and safety at 3 months in patients with primary biliary cholangitis: ENHANCE, a phase 3, randomized, placebo-controlled study. Hepatology. 2023;78:397– 415.

23. Fred RG, Steen Pedersen J, Thompson JJ, Lee J, Timshel PN, Stender S, et al. Single-cell transcriptome and cell type-specific molecular pathways of human non-alcoholic steatohepatitis. Sci. Rep. 2022;12:13484.

24. Loomba R, Amangurbanova M, Bettencourt R, Madamba E, Siddiqi H, Richards L, et al. MASH Resolution Index: development and validation of a non-invasive score to detect histological resolution of MASH. Gut. 2024;73:1343–1349.

25. Cathcart J, Barrett R, Bowness JS, Mukhopadhya A, Lynch R, Dillon JF. Accuracy of non-invasive imaging techniques for the diagnosis of MASH in patients with MASLD: A systematic review. Liver Int. 2025;45:e16127.

26. Gbadamosi SO, Evans KA, Brady BL, Hoovler A. Noninvasive tests and diagnostic pathways to MASH diagnosis in the United States: a retrospective observational study. J. Med. Econ. 2025;28:314–322.

27. Tincopa MA, Loomba R. Noninvasive tests to assess fibrosis and disease severity in metabolic dysfunction-associated steatotic liver disease. Semin. Liver Dis. 2024;44:287–299.

28. Patel K, Sebastiani G. Limitations of non-invasive tests for assessment of liver fibrosis. JHEP Rep. 2020;2:100067.

29. Koyama Y, Brenner DA. Liver inflammation and fibrosis. J. Clin. Invest. 2017;127:55–64.

30. Cholankeril G, Kramer JR, Chu J, Yu X, Balakrishnan M, Li L, et al. Longitudinal changes in fibrosis markers are associated with risk of cirrhosis and hepatocellular carcinoma in non-alcoholic fatty liver disease. J. Hepatol. 2023;78:493–500.

31. Vieira Barbosa J, Milligan S, Frick A, Broestl J, Younossi Z, Afdhal N, et al. Fibrosis-4 index can independently predict major adverse cardiovascular events in nonalcoholic fatty liver disease. Am. J. Gastroenterol. 2022;117:453–461.

32. Gbadamosi SO, Nagelhout E, Sienko D, Lamarre N, Noshad S, Ding Y, et al. Association of index and changes in fibrosis-4 score with outcomes in metabolic dysfunction-associated steatohepatitis. Gastro Hep Adv. 2025;4:100666.

33. Kamm DR, McCommis KS. Hepatic stellate cells in physiology and pathology. J. Physiol. 2022;600:1825–1837.

