## Supplemental Tables 1-3 for "Quantitative fibrosis features from digitized H&E-stained liver biopsies reveal granular insights into fibrosis and disease progression in metabolic dysfunction-associated steatohepatitis: a retrospective analysis of STELLAR-3 and STELLAR-4"

**Supplementary Tables**

**Supplementary Table S1.** Model sensitivity and positive predictive values (PPV) for model-derived and pathologist-derived cell type predictions. *indicates where the model-derived predictions pass non-inferiority test, two-sided alpha=0.05 with margin 0.1

| **Cell class** | **Model Sensitivity [95% CI]** | **Model PPV [95% CI]** | **Pathologist Sensitivity [95% CI]** | **Pathologist PPV [95% CI]** |
|---:|---:|---:|---:|---:|
| **all hepatocytes*** | 0.81 [0.77, 0.85]* | 0.83 [0.80, 0.85]* | 0.86 [0.85, 0.87] | 0.86 [0.85, 0.87] |
| normal hepatocytes* | 0.52 [0.42, 0.62] | 0.57 [0.52, 0.61]* | 0.59 [0.54, 0.62] | 0.59 [0.54, 0.62] |
| other hepatocytes* | 0.54 [0.46, 0.60]* | 0.41 [0.37, 0.45]* | 0.44 [0.40, 0.48] | 0.44 [0.40, 0.48] |
| steatotic hepatocytes* | 0.64 [0.59, 0.68]* | 0.66 [0.63, 0.71]* | 0.67 [0.64, 0.70] | 0.67 [0.64, 0.70] |
| ballooned hepatocytes | 0.54 [0.24, 0.66] | 0.52 [0.26, 0.63] | 0.67 [0.51, 0.75] | 0.67 [0.51, 0.75] |
| **all immune cells*** | 0.84 [0.81, 0.86]* | 0.39 [0.37, 0.41] | 0.54 [0.52, 0.56] | 0.54 [0.52, 0.56] |
| eosinophils | 0.54 [0.38, 0.68] | 0.45 [0.33, 0.60] | 0.62 [0.53, 0.72] | 0.62 [0.53, 0.72] |
| Neutrophils | 0.66 [0.53, 0.80]* | 0.18 [0.12, 0.25] | 0.45 [0.35, 0.55] | 0.45 [0.35, 0.55] |
| lymphocytes* | 0.63 [0.59, 0.66]* | 0.43 [0.39, 0.46] | 0.51 [0.49, 0.54] | 0.51 [0.49, 0.54] |
| plasma cells* | 0.29 [0.14, 0.43]* | 0.20 [0.11, 0.31] | 0.27 [0.15, 0.39] | 0.27 [0.15, 0.39] |
| macrophages* | 0.55 [0.51, 0.59]* | 0.16 [0.14, 0.18]* | 0.24 [0.22, 0.26] | 0.24 [0.22, 0.26] |
| **all other cells** | 0.35 [0.30, 0.40] | 0.61 [0.58, 0.65]* | 0.57 [0.54, 0.60] | 0.57 [0.54, 0.60] |
| other cells | 0.21 [0.17, 0.26] | 0.73 [0.68, 0.77]* | 0.54 [0.51, 0.57] | 0.54 [0.52, 0.57] |
| fibroblasts* | 0.55 [0.47, 0.62]* | 0.27 [0.22, 0.31] | 0.37 [0.31, 0.42] | 0.37 [0.31, 0.42] |

**Supplementary Table S2. Evaluation of zonal model performance.**

| **Component** | **Cohort** | **Model vs Consensus Correlation (ICC)** | **Average inter-pathologist correlation (ICC)** |
|---|---|---|---|
| **Portal tracts** | All slides (N=50) | 0.65 [0.49, 0.78] | 0.67 [0.59, 0.74] |
| | F-stage 0–2 (N=30) | 0.76 [0.59, 0.87] | 0.56 [0.44, 0.67] |

| | | | |
|---|---|---|---|
| | F-stage 3–4 (N=20) | 0.54 [0.29, 0.75] | 0.71 [0.55, 0.82] |
| **Central Veins** | All slides (N=50) | 0.60 [0.48, 0.71] | 0.88 [0.84, 0.91] |
| | F-stage 0–2 (N=30) | 0.58 [0.43, 0.73] | 0.84 [0.76, 0.89] |
| | F-stage 3–4 (N=20) | 0.50 [0.25, 0.69] | 0.88 [0.82, 0.92] |

**Supplementary Table S3.** Model sensitivity and positive predictive values (PPV) for model-derived and pathologist-derived fibrosis subtype predictions. *indicates where the model-derived predictions pass non-inferiority test, two-sided alpha=0.05 with margin 0.2

| **Fibrosis Subtype** | **Model Sensitivity [95% CI]** | **Model PPV [95% CI]** | **Pathologist Sensitivity [95% CI]** | **Pathologist PPV [95% CI]** |
|---|---|---|---|---|
| **pathological fibrosis*** | 0.92 [0.84, 0.96]* | 0.76 [0.65, 0.84] | 0.87 [0.80, 0.91] | 0.87 [0.80, 0.91] |
| **early-stage fibrosis*** | 0.58 [0.50, 0.64]* | 0.40 [0.32, 0.45]* | 0.49 [0.42, 0.56] | 0.49 [0.42, 0.56] |
| periportal fibrosis* | 0.14 [0.07, 0.27]* | 0.18 [0.11, 0.27]* | 0.22 [0.15, 0.31] | 0.22 [0.15, 0.31] |
| perisinusoidal fibrosis* | 0.60 [0.53, 0.66]* | 0.37 [0.28, 0.44] | 0.51 [0.43, 0.59] | 0.51 [0.43, 0.59] |
| **advanced fibrosis*** | 0.87 [0.79, 0.92]* | 0.72 [0.59, 0.81] | 0.84 [0.77, 0.89] | 0.84 [0.77, 0.89] |
| nodular fibrosis* | 0.81 [0.53, 0.94]* | 0.45 [0.25, 0.58] | 0.63 [0.44, 0.74] | 0.63 [0.44, 0.74] |
| complete septal fibrosis | 0.28 [0.15, 0.44] | 0.50 [0.33, 0.68]* | 0.55 [0.43, 0.68] | 0.55 [0.43, 0.68] |
| incomplete septal fibrosis | 0.26 [0.15, 0.43] | 0.40 [0.26, 0.54] | 0.45 [0.31, 0.59] | 0.45 [0.31, 0.59] |
| **non-pathological collagen** | 0.67 [0.45, 0.81] | 0.81 [0.66, 0.90]* | 0.85 [0.75, 0.93] | 0.85 [0.75, 0.93] |
| portal collagen | 0.50 [0.34, 0.69] | 0.79 [0.66, 0.90] | 0.78 [0.66, 0.88] | 0.78 [0.66, 0.88] |
| perivenular fibrosis | 0.59 [0.45, 0.77] | 0.50 [0.30, 0.66] | 0.65 [0.48, 0.76] | 0.65 [0.48, 0.76] |
| structural collagen | 0.74 [0.28, 0.93] | 0.80 [0.41, 0.95] | 0.85 [0.58, 0.96] | 0.85 [0.58, 0.96] |
| nonspecific collagen* | 0.28 [0.14, 0.52] | 0.09 [0.06, 0.12] | 0.37 [0.23, 0.49] | 0.37 [0.23, 0.49] |
